# Comprehensive Analysis of Juvenile Nasopharyngeal Angiofibromas via Whole Exome Sequencing

**DOI:** 10.1101/2024.05.30.24308075

**Authors:** Kiran Kumari, Shariya Afroj, Deeksha Madhry, Yash Verma, Arvind K. Kairo, Alok Thakar, Kapil Sikka, Hitesh Verma, Bhupendra Verma

## Abstract

**Objectives:** The molecular basis and mechanisms of Juvenile Nasopharyngeal Angiofibromas (JNA) pathogenesis are still unknown. Despite being a rare and benign neoplasm, JNA is a locally aggressive and potentially destructive neoplasm among head and neck tumors, typically diagnosed in young males. The advancement of genome technologies and analytical tools has provided an unparalleled opportunity to explore the intricacy of JNA. The present study provides the first evidence of the involvement of chromosome Y genes in JNA.

**Methods:** Thirteen JNA patients at an advanced disease stage and 5 age-matched male controls were registered for this study. Whole-exome sequencing (WES) analysis was conducted followed by functional analysis to understand the molecular mechanism of the JNA.

**Results:** WES analysis revealed a high prevalence of mutations in 14 genes within the protein-coding, Male-Specific Region of the Y-chromosome of young males (mean age: 13.8 ± 2.4) with JNA. These mutations, occurring at 28 distinct positions, were characterized as moderate to high impact and were prevalent in 9 JNA patients but not in the control group. The most frequently mutated genes were USP9Y and UTY, followed by KDM5D, DDX3Y and TSPY4. The expression of USP9Y, UTY and DDX3Y was found to be co-modulated, implying their coordinated regulation as a complex. Furthermore, somatic mutations were detected in genes previously linked to JNA.

**Conclusion:** The wide array of genetic and somatic mutations identified in JNA provides novel insight into JNA pathophysiology.

## INTRODUCTION

Juvenile Nasopharyngeal Angiofibroma (JNA) is a rare, benign yet rapidly expanding neoplasm found within the posterior nasal cavity and nasopharynx, posing a potential life-threatening risk due to severe hemorrhage. It constitutes roughly 0.05 to 0.5% of all head and neck tumors and affects only 1 in 150,000 individuals ^1^. Primarily affecting adolescents and prepubescent males aged 10-20 years ^1,2^, JNA displays a distinct male predominance. Very infrequently it appears in young or post-adolescent males ^3,4^. Interestingly, reports suggest a comparatively higher incidence of JNA in the Indian subcontinent compared to the Western regions ^5^. The factors underlying this increased prevalence of JNA in the Indian subcontinent remain elusive.

JNA typically presents sporadically with symptoms such as nasal obstruction and nosebleeds due to its location. Surgical resection, often through a multidisciplinary approach, is the primary treatment. Although it offers better survival outcomes, it is crucial to explore alternative approaches for approximately 37% of cases that are ineligible for surgery or exhibit residual conditions post-surgery ^6,7^.

JNA is categorized into three types based on clinical and radiological features: Type I confined to nasal structures, Type II extending into facial regions, and Type III with a sizable tumor lobe in the middle cranial fossa ^3,8^.

The exact cause of JNA remains elusive, with no hereditary pattern identified. There is a subtle increased risk in individuals with family members affected by familial adenomatous polyposis. Although predominantly occurring in adolescent boys, studies on hormonal influences in JNA have yielded inconclusive results ^2^. JNA’s molecular and genetic aspects are still evolving, with research highlighting factors contributing to the development of this rare tumor. Angiogenesis, marked by Vascular Endothelial Growth Factor (VEGF) and other angiogenic factors, is a key feature in JNA ^9,10^. Dysregulation of the Wnt/β-catenin pathway ^11–13^, chromosomal abnormalities ^14,15^, altered expression of microRNAs ^16^ and a potential association with human papillomavirus infection ^17^ have been implicated. However, comprehensive studies are crucial to better understand JNA’s etiology, molecular pathogenesis, and sex specificity due to limited current evidence.

The aim of this study was to functionally characterize the mutational spectrum in JNA pathogenesis through whole-exome sequencing and identify the male-specificity of this disease through in-depth analysis of the Y-chromosome. Here, we report the presence of multiple mutations in the Y-chromosome genes that are involved in diverse biological functions, providing a framework for future study.

## MATERIAL AND METHODS

### Patient selection

The study included 13 patients (JA_1 to JA_13) diagnosed with advanced-stage (based on Radkowski stage) Juvenile Nasopharyngeal Angiofibroma (JNA) at AIIMS, New Delhi. Five age-matched males served as controls. Clinicopathologic characteristics of JNA patients enrolled in the study are summarized in **Table 1**. Informed consent was obtained from all male participants. Blood samples collected from 2021 to 2022 were stored at -20°C. The study was conducted with approval from the Institutional Ethics Committee, AIIMS, New Delhi (IEC-494/06.08.2021, RP-31/2021), and the Institutional Biosafety Committee, AIIMS, New Delhi (IBSC 0621_BV).

**Table 1.** Summary of clinical characteristics of the study participants with JNA^#^.

|  |  |  |
| --- | --- | --- |
| **Symptoms** |  |  |
|  | Nasal bleed | 92 (12/13) |
|  | Nasal obstruction | 85 (11/13) |
|  | mass protruding through left nose | 15 (2/13) |
|  | Headache | 8 (1/13) |
|  | Cheek Swelling | 8 (1/13) |
| **Anatomical site** |  |  |
|  | Nasal cavity | 100 (13/13) |
|  | Nasopharynx | 92 (12/13) |
|  | Sinus | 62 (8/13) |
|  | Sphenopalatine foramen | 23 (3/13) |
|  | Pterygopalatine fossa | 77 (10/13) |
|  | Infratemporal fossa | 54 (7/13) |
|  | Pterygomaxillary fossa | 15 (2/13) |
| **Radkowski stage** |  |  |
|  | I | 0 (0/13) |
|  | II | 69 (9/13) |
|  | III | 31 (4/13) |
| **Sphenoid bone invasion** |  |  |
|  | No | 31 (4/13) |
|  | Yes | 69 (9/13) |
| **Preoperative embolization** |  |  |
|  | No | 23 (3/13) |
|  | Yes | 77 (10/13) |
| **Surgical approach** |  |  |
|  | Endoscopic | 69 (9/13) |
|  | Open | 31 (4/13) |
| **Intraoperative bleeding (mL)** |  |  |
|  | <500 | 31 (4/13) |
|  | 501 - 1000 | 23 (3/13) |
|  | >1001 | 46 (6/13) |
| **Recurrence** |  |  |
|  | No | 77 (10/13) |
|  | Yes | 23 (3/13) |

### Whole-exome sequencing (WES) and analysis

Whole blood was used for DNA extraction, and WES data were generated using Illumina HiSeq at Clevergene Biocorp Private Limited, Bangalore, India. Details regarding DNA sequencing, data quality control, and variant selection and annotation are available in the **Method S1**. Variants present in the coding exons were considered for screening. For further analysis, variants were filtered by removing low-impact and modifier mutations, including synonymous, intronic, non-coding transcript exon, and down-stream gene variants. High-impact and moderate-impact variants were focused, which lead to stop gain, frameshift and missense variants. In order to explore mutations in the previously reported JNA-associated genes, a gene-list was compiled by reviewing literature with generally accepted functional evidence. The details of selected genes are provided in the **Table S7**. Variants were visualized using oncoPrint of ComplexHeatmap and a karyoplot/ideogram was created using the karyoploteR package (https://www.r-project.org/) in RStudio (https://www.rstudio.com/). The human protein-protein interaction network was constructed using the STRING database (version 12).

### Real-time polymerase chain reaction (RT-PCR)

RNA was extracted from the patient’s blood, followed by DNase treatment. PCR was carried out using gene-specific primers mentioned in **Table S1**. Details are provided in the **Method S1**. U1 snRNA was used as the normalization control. Densitometric analysis was performed using ImageJ software.

### Gene ontology and pathway enrichment analyses

Gene Ontology (GO) and Kyoto Encyclopedia of Genes and Genomes (KEGG) pathway enrichment analysis was conducted using clusterProfiler implemented in the *gage* R Bioconductor package ^18^. The GO analysis included cellular components (CC), biological processes (BP), and molecular functions (MF). The GO terms and KEGG pathways with adjusted P-values ≤0.05 were considered significant for further analysis.

### Statistical Analysis

Statistical analyses were performed using the R software version 4.2.2, and statistical significance was set at a 2-tailed P-value ofLJ<LJ0.05. RT-PCR was performed in triplicates, and the statistical significance was determined using two-way ANOVA and the Graph Pad Prism software (Graph Pad Software Inc., San Diego, CA). P-value of ≤ 0.05 was considered significant. Error bars are expressed as Standard Error of the Mean (SEM).

## RESULTS

### Patient Characteristics

Thirteen patients aged between 10 and 18 years (mean age: 13.8 years) with advanced-stage JNA were analyzed alongside five male control subjects for WES. **Table 1** outlines the clinical features of patients such as tumor site, stage, surgical method, and recurrence. The study specifically focused on patients ≤ 15 years old, with most in advanced disease stages (Radkowski stage IIB-IIC for 9 patients, IIIA-IIIB for 4). Surgical preferences included 9 endoscopic and 4 open surgeries. Patient relapse was infrequent, and nearly all underwent preoperative embolization.

### WES reveals mutations in genes located in the male-specific region on the Y-chromosome

To understand the molecular pathophysiology and pathogenesis of JNA, we performed WES analysis on thirteen blood samples of JNA as compared to controls. In these samples, a total of 1133 mutations were identified on the Y-chromosome in 10 out of the 13 JNA cases. Somatic mutations were observed in the 3 patients where no mutations were detected on the Y-chromosome. These alterations were observed at 41 distinct chromosomal positions, encompassing 4 indels and 37 Single Nucleotide Polymorphisms (SNPs). These mutations comprised 1 stop gained variant, 4 frameshift variants, 23 missense variants, 1 downstream gene variant, 7 non-coding transcript exon variants, 3 intron variants, and 2 synonymous variants. Multiple mutations were found per sample across 18 genes (**Figure 1**). The genes with mutational occurrences included: AMELY, DDX3Y, KDM5D, NLGN4Y, RBMY1F, RBMY1J, RPS4Y1, RPS4Y2, TBL1Y, TSPY1, TSPY2, TSPY3, TSPY4, TSPY8, TTTY14, USP9Y, UTY and ZFY. Notably, USP9Y and UTY were the most frequently mutated genes, harboring 11 and 5 mutations respectively. It is worth mentioning that these mutations were distributed throughout the entire length of the respective genes.

**Figure 1.**
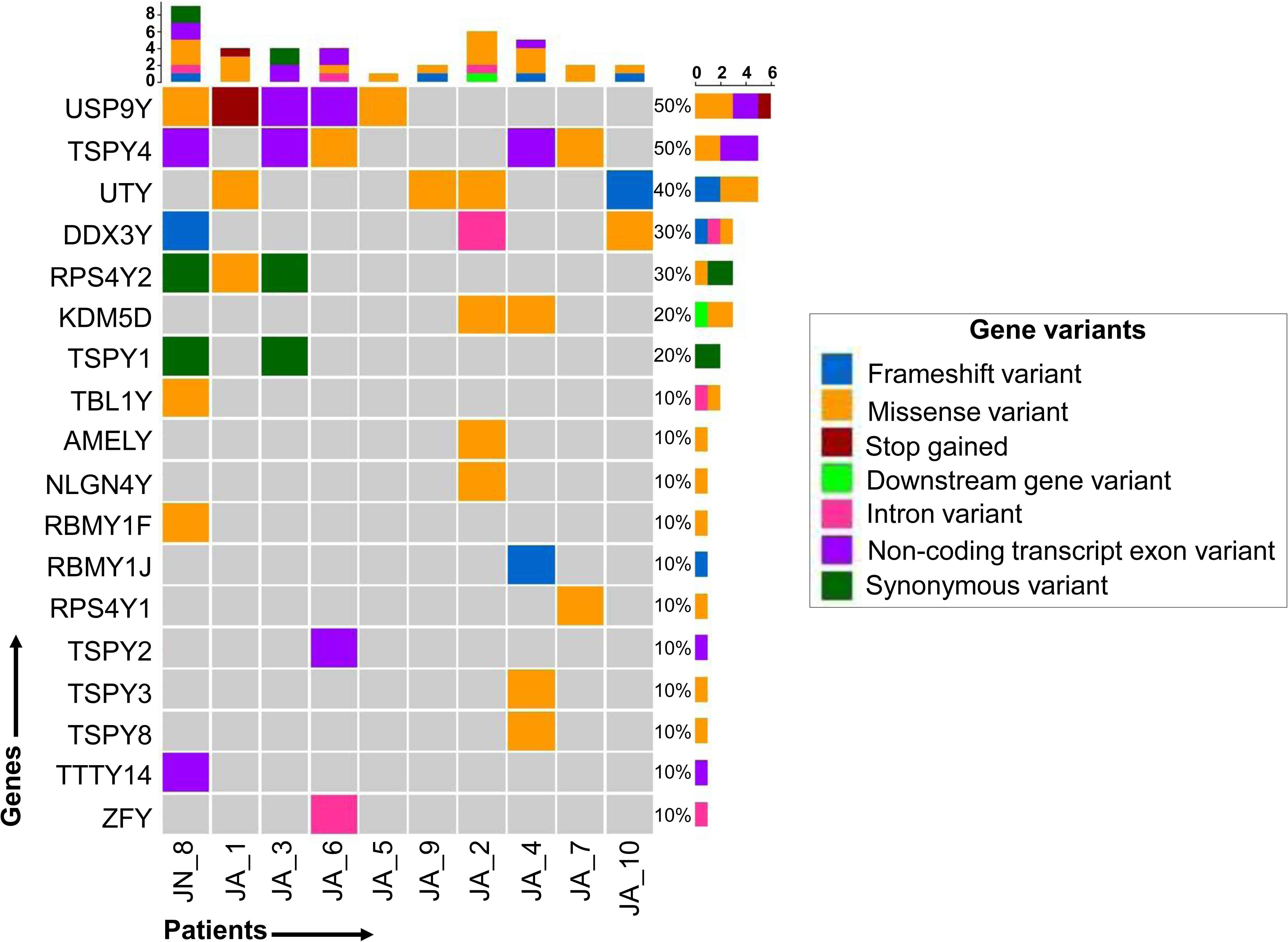
OncoPrint displays Y-chromosome germline mutations in JNA patients. Total variants per patient are above the graph. The right bar graph represents the patient percentage with at least one variant in the indicated gene.

We then focused on variants in the coding region, including stop gained, frameshift, and missense variants found at 28 distinct positions across 14 genes among the 9 patients (**Figure 2 (A), Table S2**). Notably, frequently mutated genes were USP9Y (ubiquitin specific peptidase 9 Y-linked), and UTY (ubiquitously transcribed tetratricopeptide repeat containing, Y-linked), followed by KDM5D (lysine demethylase 5D), DDX3Y (DEAD-box helicase 3 Y-linked), and TSPY4 (testis specific protein Y-linked 4). Remarkably, within the patient cohort, 3 out of 9 cases exhibited USP9Y mutations at 7 chromosomal locations, including 1 stop gained and 6 missense mutations. Similarly, UTY mutations were observed in 4 out of 9 patients across 5 chromosomal positions, involving 2 frameshift and 3 missense mutations. In DDX3Y, mutations were observed in 2 out of 9 cases at 2 chromosomal locations, including 1 frameshift and 1 missense mutation (1 upstream gene variant was identified in patient ID JA_2). Moreover, we observed a trend wherein cases at an advanced disease stage (IIIA/IIIB) exhibited a higher mutational burden of significant impact.

**Figure 2.**
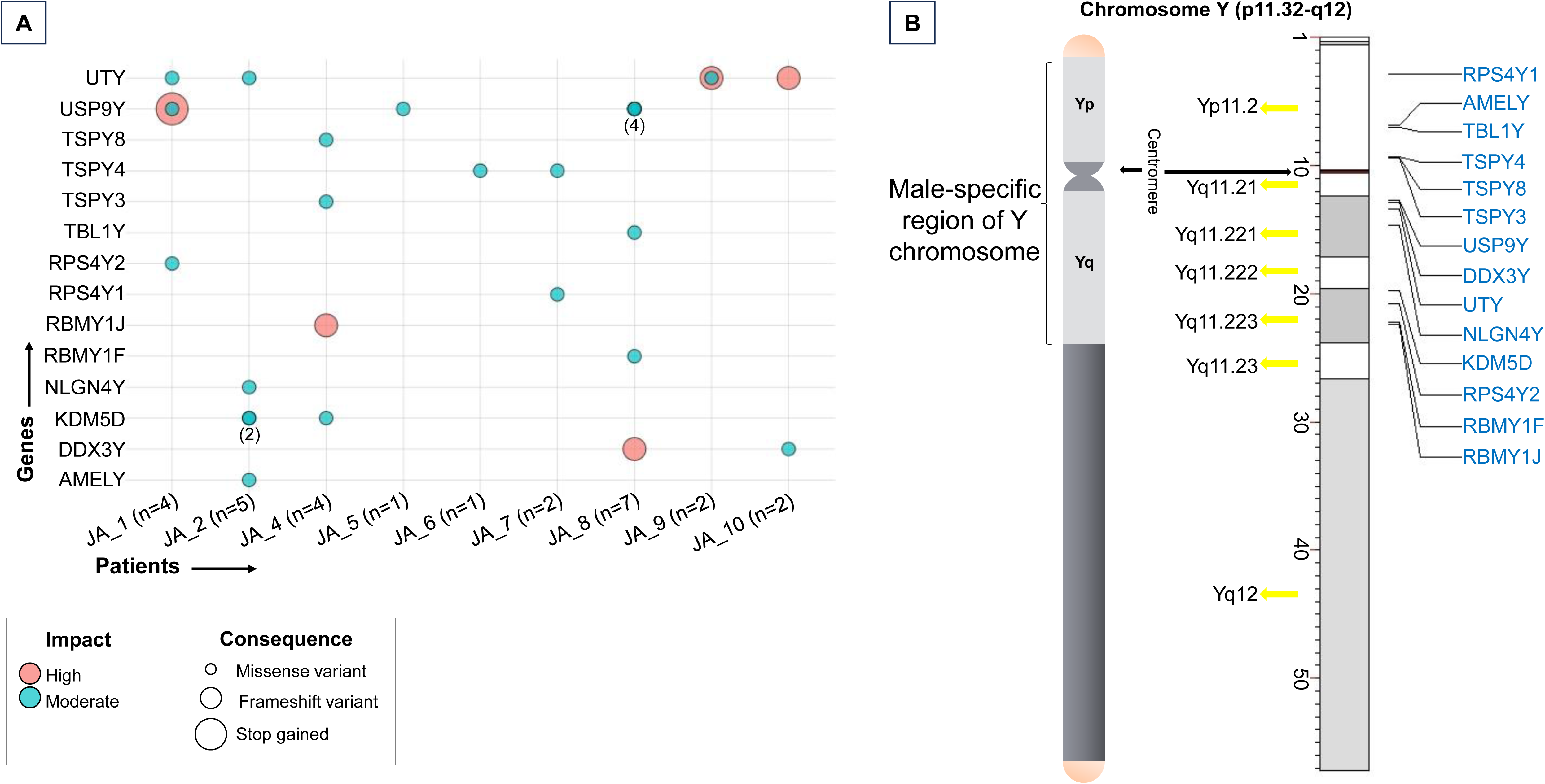
Germline mutations with potential functional impact in JNA subjects. **(A)** Dot-plot illustrates Y-chromosome gene variants in 9 patients. **(B)** Schematic and ideograms of chromosome Y showing these variants within the male-specific region of chromosome-Y.

The genomic arrangement of these 14 mutated genes on Chromosome-Y has been graphically depicted in the **Figure 2 (B)**. Mapping the genomic sequence of these mutated genes from hg38 ChrY: 1112841602 – 25241578851 revealed a notable detail– these mutations predominantly manifested within the protein-coding male-specific region of Chromosome-Y (MSY). Further investigations revealed that RPS4Y1, AMELY, TBL1Y, TSPY4, TSPY8 and TSPY3 genes resided on the Gonadoblastoma locus of Chromosome-Y, whereas USP9Y, DDX3Y and UTY genes occupied the region designated for Azoospermia factor-a. Additionally, NLGN4Y, KDM5D, RPS4Y2, RBMY1F, and RBMY1J genes were situated within the Azoospermia factor-b region of Chromosome-Y.

### JNA mutant genes modulate various cellular functions

GO and KEGG pathway analyses were conducted for functional enrichment of the 14 protein-coding mutated genes. The highly significant GO terms (adjusted p-value < 0.05) were selected for further analysis. Genes identified within the GO biological process (BP) category were predominantly associated with critical functions, including chromatin remodeling, protein demethylation, mesoderm development, protein dealkylation, gonad development, reproductive system development, demethylation, alternative mRNA splicing via spliceosome, histone modification, postsynaptic membrane assembly, and neuron cell-cell adhesion (**Figure S1**). In the GO cellular component category (CC), the genes primarily found in cytosolic small ribosomal subunit designation, indicating their involvement in fundamental cellular processes. Genes in the GO molecular function (MF) category was mainly linked to essential molecular activities such as demethylase activity, 2-oxoglutarate-dependent dioxygenase activity, histone binding, rRNA binding, structural constituent of ribosome, co-SMAD binding, and neurexin family protein binding. Notably, KEGG pathway analysis indicated a significant perturbation in the Ribosome pathway, suggesting potential disruptions in protein synthesis machinery. The details of GO terms and KEGG pathways are shown in **Table S3-S6**.

### JNA mutations lead to altered expression of USP9Y, DDX3Y, and UTY genes

Analysis with the STRING interactome database for the genes with high-impact variants revealed interactions between the USP9Y, DDX3Y, and UTY genes, forming a network (**Figure 3A**). Further investigation into these genes showed a significant modulation compared to the healthy control group. The levels of UTY, DDX3Y, and USP9Y were analyzed in the two patients (patient ID JA_10 and JA_2) through RT-PCR analysis. Patient JA_10 had one frameshift mutation (high impact) in the UTY gene and one SNP mutation (moderate impact) in the DDX3Y gene. Patient JA_2 had two SNP mutations, one in DDX3Y (modifier impact) and another in UTY genes (moderate impact). RT-PCR analysis suggested co-regulated modulation in the expression of UTY, DDX3Y and USP9Y genes. As a result of mutations in the UTY and DDX3Y genes, all three genes (UTY, DDX3Y, and USP9Y) were found to be downregulated in patient JA_10 (**Figure 3B & C**), while in patient JA_2 (**Figure 3D & E**), these genes were upregulated due to mutations in the same genes.

**Figure 3.**
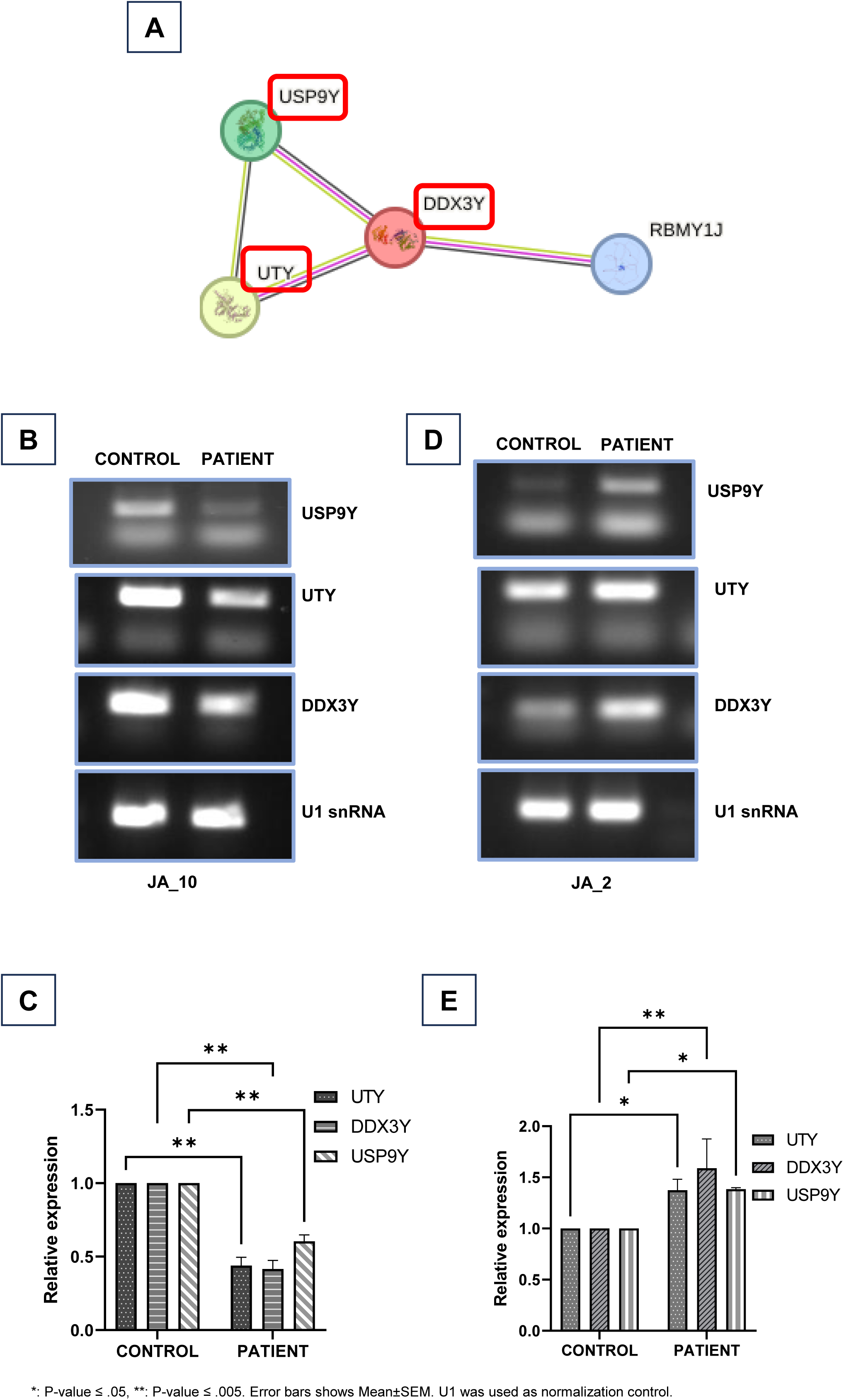
Functional analysis of USP9Y, UTY, and DDX3Y genes. **(A)** STRING interactome revealed gene interactions. RT-PCR and densitometric analysis showing expression of genes in patient JA_10 **(B and C)** and JA_2 **(D and E)**.

### Analysis of mutations in genes previously associated with JNA

We explored the mutational spectrum of genes previously associated with the molecular and somatic facets of JNA. This comprehensive panel is composed of 37 genes, categorized into five distinct groups: (i) steroid hormones and receptors such as AR, ESR and PR; (ii) growth, proliferation and tissue remodeling factors and markers including VEGF, VEGFR, APC, E-cadherin, VWF and MK167; (iii) genetic targets pertinent to JNA, such as AURKA, CTCFL, GSTM1, k-ras, c-Myc, SYK and MDM2; (iv) molecular targets encompassing c-Myc, SDC2, TLR-3, TLR-9, and TNC; and (v) Genes associated with FAP, Beta-Catenin, and the Wnt Signaling Pathway such as APC, AR, AXIN2, BMP4, E-cadherin, CTNNB1 and KDR. Upon rigorous data analysis, a spectrum of gene alterations emerged, manifesting in one or more cases of JNA (**Figure 4; Table S7**). In our scrutiny, we identified a total of 13 indels and 104 SNPs distributed across 117 distinct chromosomal positions among the 13 JNA patients. These alterations included both synonymous and non-synonymous mutations within the gene set. VWF had the highest frequency of mutations, followed by AXIN2, ESR2, CD34, MYC, APC, MKI67, TSHZ1, SYK, and TNC. After excluding non-synonymous mutations, the majority of the remaining mutations were missense alterations. Of particular interest, within MMP9, we identified a start loss in the sole identified transcript, indicating a loss of this protein function. Additionally, a stop loss mutation was observed in the SPARC gene. This comprehensive exploration of gene alterations enriches our understanding of JNA and sheds light on potential avenues for further investigation.

**Figure 4.**
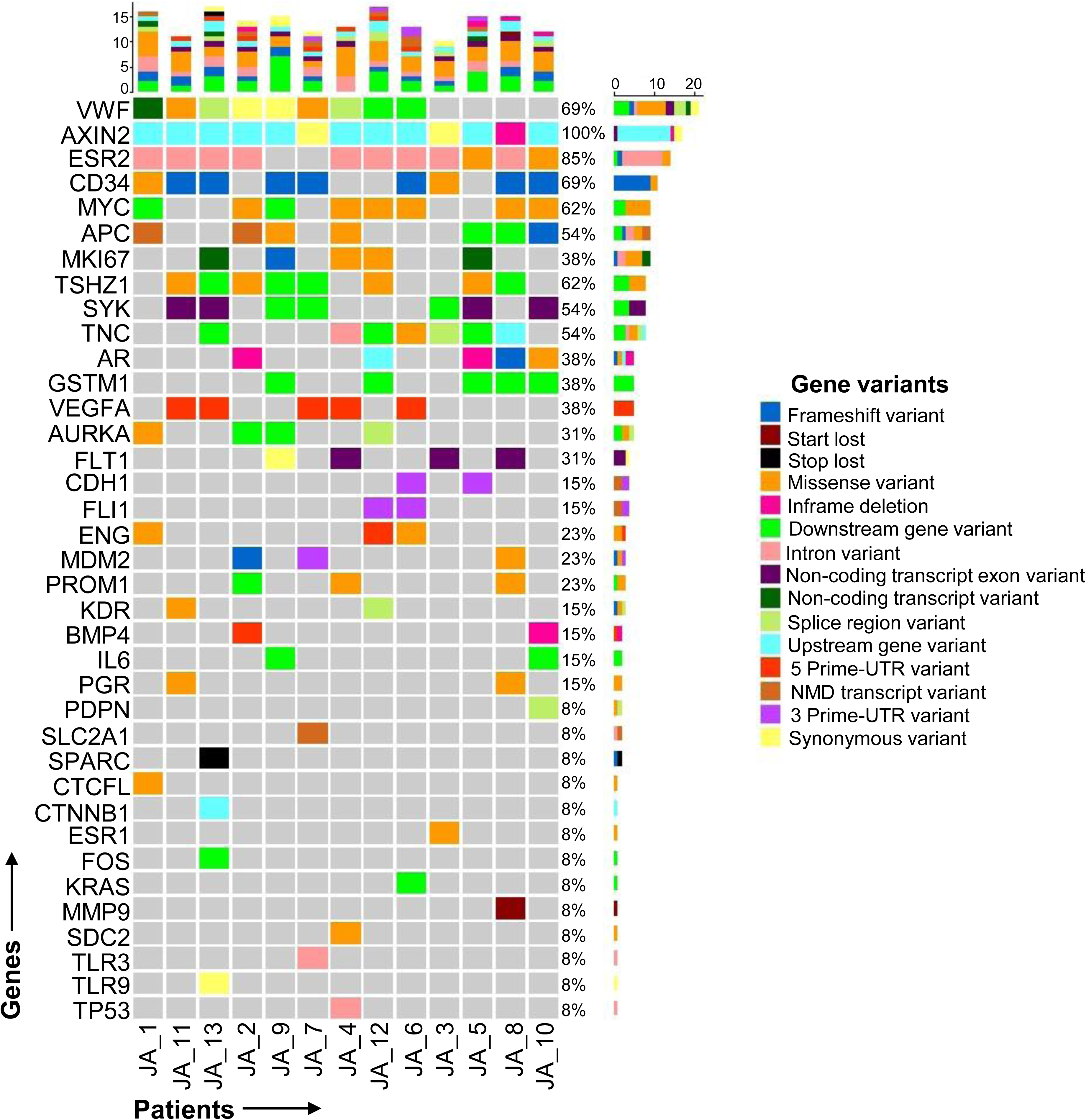
Somatic mutation spectrum of JNA. Total variants identified for individual patients is displayed above the graph. The right bar graph shows the patient percentage with at least one variant in the indicated gene.

## DISCUSSION

JNAs are aggressive vascular tumors exclusive to adolescent males, and their exact pathogenesis remains unknown. To unravel the molecular mechanism behind their male predominance, we conducted the pioneering study analyzing mutations in Y-chromosome genes within the male-specific region through exome sequencing. Additionally, we have also reported the mutations associated with genes already established to be linked with JNA.

The mutated genes, primarily within the MSY region of the Y-Chromosome, are crucial for male-specific characteristics and reproduction. Enrichment of GO terms associated with reproductive processes suggests the involvement of TSPY3, TSPY4, and TSPY8^19^ along with UTY, DDX3Y, USP9Y^20^ KDM5D, RPS4Y (RPS4Y1, RPS4Y2), and RBMY1 (RBMY1F, RBMY1J)^21,22^ in spermatogenesis and male infertility. Recent research highlights the significance of Y-chromosome genes in influencing susceptibility to various diseases, including cancers and cardiovascular diseases, with implications beyond reproductive health. Vakilian *et al*.^23^ observed increased expression of Y-linked genes (USP9Y, UTY, DDX3Y, RBMY1, and RPS4Y1) during neural cell differentiation. Elevated expression of USP9Y, DDX3Y, UTY, and RPS4Y1 in cardiovascular diseases^24^ and RPS4Y1 and DDX3Y in neurodevelopmental and neurodegenerative disorders ^23^ has been documented. These findings suggest broader functions of these genes beyond sperm production regulation.

The most frequently mutated gene identified in our JNA WES analysis is USP9Y, which encodes a deubiquitinating enzyme. USP9Y plays a crucial role in the TGF-beta/BMP signaling cascade. Our study also observed downregulation of USP9Y, suggesting NMD pathway activation to mitigate the mutational effect. USP9Y is involved in regulating protein turnover and male germ cell development^25,26^. Recently Zhu *et al*.^27^ connected USP9Y gene fusion with prostate cancer, implying that its deubiquitinating activity may impact JNA progression by modulating protein degradation pathways.

The present study identified the second most frequent mutation in UTY among the four cases examined. UTY encodes a male-specific histone demethylase responsible for the demethylation of H3K27me3 in DNA histone H3. The enrichment of GO terms under BP, such as histone lysine demethylation, histone demethylation, protein demethylation, and protein dealkylation supports this statement. Reduced expression of UTY was observed in this study. It also acts as a chaperone, and recently, deleterious missense variant was identified in the coding region of UTY genes through computational analysis ^28,29^. UTY has been reported to be involved in prostate cancer ^30^ and urothelial bladder cancer ^31^. UTY’s homology with UTX and its epigenetic functions suggest potential contributions to JNA development, particularly in terms of epigenetic modifications.

In KDM5D, a histone demethylase gene, we found three missense mutations related to epigenetic regulation. It is involved in chromosome condensation during meiosis and DNA repair. Downregulation of KDM5D is associated with increased invasiveness and metastasis in cancers ^32–34^. Its loss, particularly in males with non-small-cell lung cancer, increases mortality risk and contributes to gender-based differences ^35^. These mutations suggest a role in JNA development through epigenetic modifications.

The frameshift and missense mutation observed in DDX3Y suggest implication on different functions, as it is an RNA helicase involved in RNA metabolism. It is reported to regulates cell cycle progression in G1-S cell cycle phase via cyclin E1 ^36^. Lack of DDX3Y in males can lead to Sertoli Cell Only syndrome, impacting spermatogenesis ^37^. Downregulation of DDX3Y has been reported in hepatocellular carcinoma (HCC) ^38^ and prostate cancer ^39^, suggesting its involvement in cancer progression.

Missense or frameshift mutations were identified in several genes, including AMELY, KDM5D, NLGN4Y, RBMY1F, RBMY1J, RPS4Y1, RPS4Y2, TBL1Y, TSPY1, TSPY2, TSPY3, TSPY4, and TSPY8. NLGN4Y, implicated in synaptic functions, showed enrichment in gene ontology terms related to neural processes ^40^. Its association with autism spectrum disorders suggests a potential role of the Y-chromosome in this condition ^41,42^. RBMY1F and RBMY1J, involved in alternative mRNA splicing, have links to hepatocellular carcinoma through the Wnt/β-catenin pathway ^43,44^. RPS4Y1 and RPS4Y2, encoding ribosomal proteins, play essential roles in protein synthesis. TBL1Y, a component of the nuclear receptor corepressor complex, suggests regulatory contributions to cancer-related processes. The TSPY gene family, acting as proto-oncogenes ^45,46^ has implications in hepatocellular carcinoma and prostate cancer ^47–49^ suggesting their potential involvement in JNA development.

It is well-established that the USP9Y, UTY, and DDX3Y genes exhibit ubiquitous expression, with widespread effects beyond reproductive functions in sex organs. Maintaining dosage balance between MSY genes and their X homologs is crucial for men’s overall health ^38^. In this study, we observed co-expression of the UTY, DDX3Y, and USP9Y genes, which exhibited high-impact mutations in JNA patients. This finding aligns with the previously available reports documenting co-regulation of Chromosome-Y genes, including UTY and DDX3Y ^50^. Interestingly, we observed an opposite co-expression pattern of these three genes in two JNA patients. The downregulation of gene expression in JA_10 and upregulation in JA_2, with Radkowski stage IIIB and IIB respectively, suggest their association with increased aggressiveness of the disease. Since the Y-chromosome is unique to men, these findings underscore the potential involvement of Y-chromosome genes in JNA development, highlighting their pivotal role in influencing susceptibility. Given that this study is the first of its kind, further research is crucial to unravel the precise roles of these genes in male-specific and gender-biased JNA development, progression, aggressiveness and outcome. Continued investigation will help to determine the clinical impact of Chromosome-Y genes and contribute to improved diagnostics and therapeutic strategies for JNA.

The study categorizes previously reported genetic markers associated with JNA into five groups based on types and functions. The first group, including AR, ESR1, and PR genes, crucial in sexual development, exhibits high-impact mutations in AR ^51^, suggesting potential influences on cancer. The second group involves markers linked to growth, proliferation, and tissue remodeling, such as VEGF, VEGFR, APC, E-cadherin, VWF, and MKI67, providing insights into vasculogenesis ^52^ angiogenesis, and tumorogenesis ^53^ VEGF is linked to high-vessel density in JNA ^54^ APC mutations, common in FAP-associated JNA, affect the Wnt pathway’s involvement in JNA pathophysiology ^55^ E-cadherin mutations suppress tumors and are linked to various cancers ^56^ Notably, VWF and MKI67 mutations suggest links to JNA development as they are involved in maintaining vascular integrity and cell growth and proliferation ^10,57^. The third group includes AURKA, c-Myc, CTCFL, MDM2, GSTM1, K-RAS, and SYK genes, pivotal in cell cycle regulation ^58^ and tumor progression ^59^. The fourth group, featuring c-Myc, SDC2, TLR-3, TLR-9, and TNC genes, reveals diverse mechanisms such as regulation of cell cycle, cell adhesion and signaling, immune responses, and tissue homeostasis in JNA progression, with c-Myc and TNC showing high mutation frequencies ^12,60,61^. The fifth group connects JNA with FAP, Beta-Catenin, and the Wnt Signaling Pathway through genes like APC, AR, AXIN2, BMP4, E-cadherin, CTNNB1, and KDR, emphasizing their crucial roles and interactions ^2,11,62^. Additionally, variants in MMP9, CD34, ESR2, and TSHZ1 that are involved in the regulation of tissue development, repair, and homeostasis support their connection with JNA pathogenesis, offering potential therapeutic targets and enhancing our understanding of JNA.

## CONCLUSION

WES unveils the molecular mechanisms and sex specificity linked to JNA. Exploring mutations in the Y-chromosome male-specific region highlights the biological implications, emphasizing the significance of this chromosome in health and disease, particularly in cancer. Notably, specific mutation in UTY, USP9 and DDX3Y genes leads to their mutual alterations or co-regulated gene expression as observed in JNA patients, indicative of their involvement in JNA aggressiveness. Meanwhile, mutations in various genes like VEGF, APC, E-cadherin, and more suggest a diverse range of genetic factors contributing to JNA. This research marks a promising avenue for understanding JNA pathogenesis, paving the way for improved prevention, prognosis, and treatment strategies.

## STATEMENTS & DECLARATIONS

### AUTHOR CONTRIBUTIONS

Dr. Bhupendra Verma and Dr. Hitesh Verma had full access to all the data in the study and take responsibility for the integrity of the data and the accuracy of the data analysis. Dr. Kiran Kumari and Shariya Afroj contributed equally to this study.

Concept and design: Dr. Bhupendra Verma and Dr. Hitesh Verma.

Acquisition, analysis, or interpretation of data: Dr. Kiran Kumari, Shariya Afroj, Deeksha Madhry, Dr. Bhupendra Verma and Dr. Hitesh Verma.

Drafting of the manuscript: Dr. Kiran Kumari, Shariya Afroj, Deeksha Madhry, Dr. Bhupendra Verma and Dr. Hitesh Verma.

Critical revision of the manuscript for important intellectual content: All authors. Statistical analysis: Dr. Kiran Kumari, Shariya Afroj and Dr. Bhupendra Verma Obtained funding: Dr. Bhupendra Verma and Dr. Hitesh Verma.

Administrative, technical, or material support: Dr. Bhupendra Verma and Dr. Hitesh Verma. Supervision: Dr. Bhupendra Verma and Dr. Hitesh Verma.

## ACKNOWLEDGEMENTS

The authors are grateful to the patients and their families, as well as clinical and non-clinical staff at All India Institute of Medical Sciences, Ansari Nagar, New Delhi for their participation in the study. The authors also thank team from Clevergene Biocorp Pvt. Ltd., Bengaluru, India, for providing assistance with whole-exome sequencing. No compensation was received.

## DATA AVAILABILITY

All data and materials described in this publication are included in the manuscript and additional supporting files. Raw datasets used and/or analyzed during the current study will be made available upon request to the corresponding authors.

## ETHICS APPROVAL

The study was conducted with approval from the Institutional Ethics Committee, AIIMS, New Delhi (IEC-494/06.08.2021, RP-31/2021), and the Institutional Biosafety Committee, AIIMS, New Delhi (IBSC 0621_BV).

## CONSENT TO PARTICIPATE

Informed consent was obtained from all individual participants included in the study.

## SUPPLEMENTARY INFORMATION

The online version contains supplementary material.

