## Supplementary File for "Comprehensive Analysis of Juvenile Nasopharyngeal Angiofibromas via Whole Exome Sequencing"

**Contents:**

**Method S1.** Detailed Methodology

**Table S1.** List of Primers used in this study

**Table S2.** The list of the identified mutations in the Y-chromosome of JNA patients

**Table S3.** Significantly enriched GO terms of Biological Process in Y chromosome mutated genes

**Table S4.** Significantly enriched GO terms of Molecular Function in Y chromosome mutated genes

**Table S5.** Significantly enriched GO terms of Cellular Component in Y chromosome mutated genes

**Table S6.** Significantly regulated KEGG pathways of genes mutated in Y chromosome

**Table S7.** Mutational spectrum of the previous reported genes in context of JNA

**Figure S1.** Plot showing the Y chromosome mutated genes involved in GO terms of Biological Process

**Supplementary References**

**Method S1. Detailed Methodology**

**Whole-exome sequencing (WES) and analysis**

Whole blood was used for DNA extraction using QIAGEN Blood kit according to the manufacturer’s protocol. WES data was generated using Illumina HiSeq at Clevergene Biocorp Private Limited, Bangalore, India. Data quality was checked using FastQC and MultiQC software. The data was checked for base call quality distribution, % bases above Q20, Q30, %GC, and sequencing adapter contamination. All the samples have passed the QC threshold (Q20>95%). Raw sequence reads were processed to remove adapter sequences and low-quality bases using fastp. The quality trimmed reads were aligned to Human reference genome build GRCh38 using bwa mem algorithm. The alignments were processed to remove PCR duplicates using PICARD tools. The base qualities were recalibrated, and variants were called using GATK haplotype caller pipeline. Variants were recalibrated as per the GATK best practice guidelines. The variants were annotated using Ensembl Variant Effect Predictor (VEP). All transcripts (Ensembl/GENCODE,Refseq) were considered for consequence estimation. The variants were annotated with no selection (no filter) option in VEP. The vcf files were processed using custom scripts to convert the data in tab delimited text format files. The CSV file was splitted based on chromosomes for easier visualization of the data.

**Real-time polymerase chain reaction (RT-PCR)**

RNA was extracted using the NucleoSpin RNA Blood kit (NucleoSpin, MACHEREY-NAGEL, Duren, Germany) according to the manufacturer’s protocol. Following DNase treatment, 250ng of RNA was reverse transcribed to cDNA using MLV-RT (Promega). PCR experiments were carried out using the GoTaq Flexi DNA polymerase kit (Promega) according to the manufacturer’s protocol. Specific primers were designed using the Primer3 software mentioned in **eTable 1**. Cycling conditions were: initial denaturation hold at 98°C for 3min, denaturation at 98°C for 30sec, annealing at 52-58°C for 20sec, extension at 72°C for 30sec, final extension at 72°C for 5min, amplification (34 cycle). Soak at 4°C infinite. U1 was used as the normalization control. The densitometric analysis performed using ImageJ software.

**Table S1. List of Primers used in this study**

| **Gene** | **Sequence** | **Product size** |
| --- | --- | --- |
| U1 Sn RNA | FP: 5’TAAGCACTCGAGATACTTACCTGGCAG 3’ | 150 bp |
|  | RP: 5’TAAGCATCTAGACAGGGGAAAGCGCGAA 3’ |  |
| USP9Y | FP: 5’CTTTGTGATCGTGAAGCCTG 3’ | 187 bp |
|  | RP: 5’TCCTTTCTCGACAATTGACAGC 3’ |  |
| UTY | FP: 5’ATCTAGCGTCTCTCAGCCTG 3’ | 242 bp |
|  | RP: 5’ GCTCTTCTGTGCTGCTGTTC 3’ |  |
| DDX3Y | FP: 5’ ACTTTAGTGTTTGTGGAGACCA 3’ | 139 bp |
|  | RP: 5’ CTGAGCGAAACTGGTGAAGG 3’ |  |

**Table S2. The list of the identified mutations in the Y-chromosome of JNA patients**

| **Gene** | **Gene description** | **Type** | **Impact** | **Consequence** | **Patient id** |
| --- | --- | --- | --- | --- | --- |
| AMELY (n=1) | Amelogenin Y-Linked | SNP | MODERATE | Missense variant | JA_2 (n=1) |
| DDX3Y (n=2) | DEAD-Box Helicase 3 Y-Linked | INDEL | HIGH | Frameshift variant | JA_8 (n=1) |
|  |  | SNP | MODERATE | Missense variant | JA_10 (n=1) |
| KDM5D (n=3) | Lysine Demethylase 5D | SNP | MODERATE | Missense variant | JA_2 (n=2) |
|  |  |  |  |  | JA_4 (n=1) |
| NLGN4Y (n=1) | Neuroligin-4, Y-linked | SNP | MODERATE | Missense variant | JA_2 (n=1) |
| RBMY1F (n=1) | RNA Binding Motif Protein Y-Linked Family 1 Member F | SNP | MODERATE | Missense variant | JA_8 (n=1) |
| RBMY1J (n=1) | RNA Binding Motif Protein Y-Linked Family 1 Member J | INDEL | HIGH | Frameshift variant | JA_4 (n=1) |
| RPS4Y1 (n=1) | Ribosomal Protein S4, Y-Linked 1 | SNP | MODERATE | Missense variant | JA_7 (n=1) |
| RPS4Y2 (n=1) | Ribosomal Protein S4, Y-Linked 2 | SNP | MODERATE | Missense variant | JA_1 (n=1) |
| TBL1Y (n=1) | Transducin Beta Like 1 Y-Linked | SNP | MODERATE | Missense variant | JA_8 (n=1) |
| TSPY3 (n=1) | Testis Specific Protein Y-Linked 3 | SNP | MODERATE | Missense variant | JA_4 (n=1) |
| TSPY4 (n=2) | Testis Specific Protein Y-Linked 4 | SNP | MODERATE | Missense variant | JA_7 (n=1) |
|  |  |  |  |  | JA_6 (n=1) |
| TSPY8 (n=1) | Testis Specific Protein Y-Linked 8 | SNP | MODERATE | Missense variant | JA_4 (n=1) |
| USP9Y (n=7) | Ubiquitin specific peptidase 9, Y-linked | SNP (n=7) | HIGH (n=1) | Stop gained (n=1) | JA_1 (n=1) |
|  |  |  | MODERATE (n=6) | Missense variant (n=6) | JA_8 (n=4) |
|  |  |  |  |  | JA_5 (n=1) |
|  |  |  |  |  | JA_1 (n=1) |
| UTY (n=5) | Ubiquitously Transcribed Tetratricopeptide Repeat Containing, Y-Linked | INDEL (n=2) | HIGH (n=2) | Frameshift variant (n=2) | JA_10 (n=1) |
|  |  |  |  |  | JA_9 (n=1) |
|  |  | SNP (n=3) | MODERATE (n=3) | Missense variant (n=3) | JA_9 (n=1) |
|  |  |  |  |  | JA_2 (n=1) |
|  |  |  |  |  | JA_1 (n=1) |

n=number of variants identified

**Table S3. Significantly enriched GO terms of Biological Process in Y chromosome mutated genes**

| **ID** | **Description** | **P-value** | **Adjusted**  **P-value** | **Gene ID** | **Count** |
| --- | --- | --- | --- | --- | --- |
| GO:0006334 | nucleosome assembly | 8.85E-05 | 0.002588 | TSPY3/TSPY4/TSPY8 | 3 |
| GO:0007498 | mesoderm development | 0.000125 | 0.002588 | TSPY3/TSPY4/TSPY8 | 3 |
| GO:0006338 | chromatin remodeling | 0.000128 | 0.002588 | KDM5D/TSPY3/TSPY4/TSPY8 | 4 |
| GO:0034728 | nucleosome organization | 0.000188 | 0.002588 | TSPY3/TSPY4/TSPY8 | 3 |
| GO:0070076 | histone lysine demethylation | 0.00019 | 0.002588 | KDM5D/UTY | 2 |
| GO:0016577 | histone demethylation | 0.000204 | 0.002588 | KDM5D/UTY | 2 |
| GO:0006482 | protein demethylation | 0.000249 | 0.002588 | KDM5D/UTY | 2 |
| GO:0008214 | protein dealkylation | 0.000249 | 0.002588 | KDM5D/UTY | 2 |
| GO:0008406 | gonad development | 0.000609 | 0.005128 | TSPY3/TSPY4/TSPY8 | 3 |
| GO:0045137 | development of primary sexual characteristics | 0.000647 | 0.005128 | TSPY3/TSPY4/TSPY8 | 3 |
| GO:0065004 | protein-DNA complex assembly | 0.00068 | 0.005128 | TSPY3/TSPY4/TSPY8 | 3 |
| GO:0071824 | protein-DNA complex subunit organization | 0.001005 | 0.006953 | TSPY3/TSPY4/TSPY8 | 3 |
| GO:0007548 | sex differentiation | 0.001125 | 0.006979 | TSPY3/TSPY4/TSPY8 | 3 |
| GO:0070988 | demethylation | 0.001335 | 0.006979 | KDM5D/UTY | 2 |
| GO:0048608 | reproductive structure development | 0.001339 | 0.006979 | TSPY3/TSPY4/TSPY8 | 3 |
| GO:0061458 | reproductive system development | 0.00139 | 0.006979 | TSPY3/TSPY4/TSPY8 | 3 |
| GO:0060485 | mesenchyme development | 0.001429 | 0.006979 | TSPY3/TSPY4/TSPY8 | 3 |
| GO:0000380 | alternative mRNA splicing, via spliceosome | 0.001596 | 0.00736 | RBMY1F/RBMY1J | 2 |
| GO:0016570 | histone modification | 0.004641 | 0.020273 | KDM5D/TBL1Y/UTY | 3 |
| GO:0097090 | presynaptic membrane organization | 0.007383 | 0.029182 | NLGN4Y | 1 |
| GO:0097104 | postsynaptic membrane assembly | 0.007383 | 0.029182 | NLGN4Y | 1 |
| GO:0007158 | neuron cell-cell adhesion | 0.011789 | 0.044477 | NLGN4Y | 1 |

**Table S4. Significantly enriched GO terms of Molecular Function in Y chromosome mutated genes**

| **ID** | **Description** | **P-value** | **Adjusted**  **P-value** | **Gene ID** | **Count** |
| --- | --- | --- | --- | --- | --- |
| GO:0032452 | histone demethylase activity | 0.000230229 | 0.003683669 | KDM5D/UTY | 2 |
| GO:0140457 | protein demethylase activity | 0.000230229 | 0.003683669 | KDM5D/UTY | 2 |
| GO:0032451 | demethylase activity | 0.000411037 | 0.004384398 | KDM5D/UTY | 2 |
| GO:0042393 | histone binding | 0.000880192 | 0.005917949 | TSPY3/TSPY4/TSPY8 | 3 |
| GO:0016706 | 2-oxoglutarate-dependent dioxygenase activity | 0.00092468 | 0.005917949 | KDM5D/UTY | 2 |
| GO:0019843 | rRNA binding | 0.001151564 | 0.006141673 | RPS4Y1/RPS4Y2 | 2 |
| GO:0051213 | dioxygenase activity | 0.002203144 | 0.010071515 | KDM5D/UTY | 2 |
| GO:0003735 | structural constituent of ribosome | 0.00765072 | 0.026642006 | RPS4Y1/RPS4Y2 | 2 |
| GO:0016705 | oxidoreductase activity, acting on paired donors, with incorporation or reduction of molecular oxygen | 0.007904245 | 0.026642006 | KDM5D/UTY | 2 |
| GO:0070410 | co-SMAD binding | 0.008325627 | 0.026642006 | USP9Y | 1 |
| GO:0042043 | neurexin family protein binding | 0.010585055 | 0.030792889 | NLGN4Y | 1 |
| GO:0030021 | extracellular matrix structural constituent conferring compression resistance | 0.01658685 | 0.044231601 | AMELY | 1 |

**Table S5. Significantly enriched GO terms of Cellular Component in Y chromosome mutated genes**

| **ID** | **Description** | **P-value** | **Adjusted**  **P-value** | **Gene ID** | **Count** |
| --- | --- | --- | --- | --- | --- |
| GO:0022627 | cytosolic small ribosomal subunit | 0.000448598 | 0.017046713 | RPS4Y1/RPS4Y2 | 2 |
| GO:0015935 | small ribosomal subunit | 0.00130889 | 0.024868907 | RPS4Y1/RPS4Y2 | 2 |
| GO:0022626 | cytosolic ribosome | 0.002415068 | 0.030590866 | RPS4Y1/RPS4Y2 | 2 |

**Table S6: Significantly regulated KEGG pathways of genes mutated in Y chromosome**

| **ID** | **Description** | **P-value** | **Adjusted**  **P-value** | **Gene ID** | **Count** |
| --- | --- | --- | --- | --- | --- |
| hsa03010 | Ribosome | 0.002203687 | 0.008433468 | RPS4Y1/RPS4Y2 | 2 |
| hsa05171 | Coronavirus disease - COVID-19 | 0.004216734 | 0.008433468 | RPS4Y1/RPS4Y2 | 2 |

**Table S7: Mutational spectrum of the previous reported genes in context of JNA**

| **Gene ^(Reference*)^** | **Gene description** | **TYPE** | **IMPACT** | **Consequence** | **Patient id** |
| --- | --- | --- | --- | --- | --- |
| APC ^1,2^ | Adenomatous polyposis coli | INDEL | HIGH | frameshift_variant | JA_10 |
|  |  | SNP | MODIFIER | intron_variant&NMD_transcript_variant | JA_1 JA_2 |
|  |  | SNP | MODERATE | missense_variant | JA_9 |
|  |  | SNP | MODERATE | missense_variant | JA_4 |
|  |  | SNP | MODIFIER | downstream_gene_variant | JA_5 |
|  |  | SNP | MODIFIER | downstream_gene_variant | JA_8 |
| AR ^3^ | Androgen receptor | INDEL | MODERATE | inframe_deletion | JA_2 JA_5 |
|  |  | SNP | MODIFIER | upstream_gene_variant | JA_12 |
|  |  | SNP | MODERATE | missense_variant | JA_10 |
|  |  | INDEL | HIGH | frameshift_variant | JA_8 |
| AURKA ^4^ | Aurora kinase A | SNP | MODIFIER | downstream_gene_variant | JA_2 JA_9 |
|  |  | SNP | MODERATE | missense_variant | JA_1 |
|  |  | SNP | MODERATE | missense_variant&splice_region_variant | JA_12 |
| AXIN2 ^5^ | AXIS inhibition protein 2 | SNP | MODIFIER | non_coding_transcript_exon_variant | JA_7 |
|  |  | INDEL | MODERATE | inframe_deletion | JA_8 |
|  |  | SNP | MODIFIER | upstream_gene_variant | JA_1 JA_2 JA_3 JA_4 JA_5 JA_11 JA_6 JA_12 JA_7 JA_8 JA_9 JA_10 JA_13 |
|  |  | SNP | LOW | synonymous_variant | JA_3 JA_7 |
| BMP4 ^1^ | Bone morphogenetic protein 4 | INDEL | MODERATE | inframe_deletion | JA_10 |
|  |  | SNP | MODIFIER | 5_prime_UTR_variant | JA_2 |
| CD34 ^6^ | Hematopoietic Progenitor Cell Antigen CD34 | SNP | MODERATE | missense_variant | JA_1 |
|  |  | INDEL | HIGH | frameshift_variant | JA_1 JA_3 JA_11 JA_6 JA_7 JA_8 JA_9 JA_10 JA_13 |
|  |  | SNP | MODERATE | missense_variant | JA_3 |
| CDH1^7,8^ | Cadherin 1 | SNP | MODIFIER | 3_prime_UTR_variant&NMD_transcript_variant | JA_5 |
|  |  | SNP | MODIFIER | 3_prime_UTR_variant&NMD_transcript_variant | JA_6 |
| CTCFL ^9^ | CCCTC-Binding Factor Like | SNP | MODERATE | missense_variant | JA_1 |
| CTNNB1 ^10^ | Catenin Beta 1 | SNP | MODIFIER | upstream_gene_variant | JA_13 |
| ENG ^6,11^ | Endoglin | SNP | MODERATE | missense_variant | JA_1 |
|  |  | SNP | MODERATE | missense_variant | JA_6 |
|  |  | SNP | MODIFIER | 5_prime_UTR_variant | JA_12 |
| ESR1 ^2^ | Estrogen Receptor 1 | SNP | MODERATE | missense_variant | JA_3 |
| ESR2 ^2^ | Estrogen receptor 2 | SNP | MODIFIER | intron_variant | JA_1 JA_12 JA_13 |
|  |  | SNP | MODIFIER | intron_variant | JA_5 |
|  |  | SNP | MODIFIER | intron_variant | JA_8 |
|  |  | SNP | MODIFIER | intron_variant | JA_4 |
|  |  | SNP | MODIFIER | intron_variant | JA_6 |
|  |  | SNP | MODIFIER | intron_variant | JA_6 |
|  |  | SNP | MODIFIER | intron_variant | JA_6 |
|  |  | SNP | MODIFIER | intron_variant | JA_2 |
|  |  | SNP | MODIFIER | intron_variant | JA_3 |
|  |  | SNP | MODIFIER | downstream_gene_variant | JA_1 |
|  |  | SNP | MODERATE | missense_variant | JA_10 |
|  |  | INDEL | HIGH | frameshift_variant | JA_11 |
|  |  | SNP | MODIFIER | intron_variant | JA_11 JA_12 |
|  |  | SNP | MODERATE | missense_variant | JA_5 |
| FLI1 ^2,6^ | Friend leukemia integration 1 transcription factor | SNP | MODIFIER | 3_prime_UTR_variant&NMD_transcript_variant | JA_6 JA_12 |
| FLT1 ^11^ | Fms-Related Tyrosine Kinase 1 (Vascular Endothelial Growth Factor/Vascular Permeability Factor Receptor) | SNP | MODIFIER | non_coding_transcript_exon_variant | JA_3 JA_4 JA_8 |
|  |  | SNP | LOW | synonymous_variant | JA_9 |
| FOS ^11^ | Fos Proto-Oncogene | SNP | MODIFIER | downstream_gene_variant | JA_13 |
| GSTM1 ^1^ | Glutathione S-transferase Mu 1 | SNP | MODIFIER | downstream_gene_variant | JA_9 |
|  |  | SNP | MODIFIER | downstream_gene_variant | JA_8 |
|  |  | SNP | MODIFIER | downstream_gene_variant | JA_5 JA_12 JA_8 JA_10 |
| IL6 ^12^ | Interleukin 6 | SNP | MODIFIER | downstream_gene_variant | JA_9 JA_10 |
| KDR ^1^ | Kinase insert domain receptor | INDEL | HIGH | frameshift_variant&splice_region_variant | JA_12 |
|  |  | SNP | MODERATE | missense_variant | JA_11 |
| KRAS ^1^ | Kirsten rat sarcoma virus | SNP | MODIFIER | downstream_gene_variant | JA_6 |
| MDM2 ^1^ | Murine double minute 2 | SNP | MODERATE | missense_variant | JA_8 |
|  |  | INDEL | HIGH | frameshift_variant | JA_2 |
|  |  | SNP | MODIFIER | 3_prime_UTR_variant | JA_7 |
| MKI67 ^13^ | Marker Of Proliferation Ki-67 | SNP | MODERATE | missense_variant | JA_4 |
|  |  | SNP | MODERATE | missense_variant | JA_13 |
|  |  | SNP | MODERATE | missense_variant | JA_4 |
|  |  | SNP | MODERATE | missense_variant | JA_5 JA_13 |
|  |  | INDEL | HIGH | frameshift_variant | JA_9 |
|  |  | SNP | MODERATE | missense_variant | JA_5 JA_13 |
|  |  | SNP | MODERATE | missense_variant | JA_5 JA_13 |
|  |  | SNP | MODERATE | missense_variant | JA_5 JA_13 |
|  |  | SNP | MODERATE | missense_variant | JA_5 JA_13 JA_12 |
|  |  | SNP | MODERATE | missense_variant | JA_4 |
|  |  | SNP | MODIFIER | intron_variant&non_coding_transcript_variant | JA_5 JA_13 |
| MMP9 ^11^ | Matrix metallopeptidase 9 | SNP | HIGH | start_lost | JA_8 |
| MYC ^14^ | Myc myelocytomatosis oncogene | SNP | MODERATE | missense_variant | JA_2 JA_4 JA_8 JA_10 |
|  |  | SNP | MODERATE | missense_variant | JA_6 JA_12 |
|  |  | SNP | MODIFIER | downstream_gene_variant | JA_1 JA_12 JA_9 |
| PDPN ^2,6^ | Podoplanin | SNP | MODERATE | missense_variant&splice_region_variant | JA_10 |
| PGR ^1^ | Progesterone Receptor | SNP | MODERATE | missense_variant | JA_8 |
|  |  | SNP | MODERATE | missense_variant | JA_11 |
| PROM1 ^15^ | Prominin 1 | SNP | MODIFIER | downstream_gene_variant | JA_2 |
|  |  | SNP | MODERATE | missense_variant | JA_8 |
|  |  | SNP | MODERATE | missense_variant | JA_4 |
|  |  | SNP | MODERATE | missense_variant | JA_4 |
|  |  | SNP | MODERATE | missense_variant | JA_4 |
| SDC2 ^16^ | Syndecan-2 | SNP | MODERATE | missense_variant | JA_4 |
| SLC2A1 ^2,6^ | Solute carrier family 2 member 1 | SNP | MODIFIER | intron_variant&NMD_transcript_variant | JA_7 |
| SPARC ^2,6^ | Secreted Protein Acidic And Cysteine Rich | INDEL | HIGH | frameshift_variant&stop_lost | JA_13 |
| SYK ^14^ | Spleen associated tyrosine kinase | SNP | MODIFIER | non_coding_transcript_exon_variant | JA_5 JA_11 JA_10 JA_13 |
|  |  | SNP | MODIFIER | downstream_gene_variant | JA_3 JA_7 JA_9 |
|  |  | SNP | MODIFIER | downstream_gene_variant | JA_5 |
| TLR3 ^2,17^ | Toll like receptor 3 | SNP | MODIFIER | intron_variant | JA_7 |
| TLR9 ^2,17^ | Toll like receptor 9 | SNP | LOW | synonymous_variant | JA_13 |
| TNC ^18^ | Tenascin-C | SNP | MODERATE | missense_variant&splice_region_variant | JA_3 |
|  |  | SNP | MODIFIER | downstream_gene_variant | JA_13 |
|  |  | SNP | MODIFIER | downstream_gene_variant | JA_5 JA_12 |
|  |  | SNP | MODIFIER | intron_variant | JA_4 |
|  |  | SNP | MODERATE | missense_variant | JA_6 |
|  |  | SNP | MODIFIER | upstream_gene_variant | JA_8 |
| TP53 ^12^ | Tumor protein p53 | SNP | MODIFIER | intron_variant | JA_4 |
| TSHZ1 ^19^ | Teashirt zinc finger homeobox 1 | SNP | MODERATE | missense_variant | JA_5 |
|  |  | SNP | MODIFIER | downstream_gene_variant | JA_7 JA_8 JA_9 JA_13 |
|  |  | SNP | MODERATE | missense_variant | JA_2 JA_11 |
|  |  | SNP | MODERATE | missense_variant | JA_12 |
| VEGFA ^6,20^ | Vascular endothelial growth factor A | SNP | MODIFIER | 5_prime_UTR_variant | JA_4 JA_11 JA_6 JA_7 JA_13 |
| VWF ^6^ | Von Willebrand factor | SNP | MODERATE | missense_variant&splice_region_variant | JA_1 JA_4 JA_13 |
|  |  | SNP | LOW | synonymous_variant | JA_2 JA_9 |
|  |  | INDEL | HIGH | frameshift_variant | JA_1 |
|  |  | SNP | MODERATE | missense_variant | JA_1 |
|  |  | SNP | MODIFIER | intron_variant&non_coding_transcript_variant | JA_1 |
|  |  | INDEL | HIGH | frameshift_variant | JA_1 |
|  |  | SNP | MODIFIER | downstream_gene_variant | JA_9 |
|  |  | SNP | MODIFIER | downstream_gene_variant | JA_11 JA_6 JA_12 |
|  |  | SNP | MODERATE | missense_variant | JA_11 |
|  |  | SNP | MODIFIER | downstream_gene_variant | JA_11 JA_6 |
|  |  | SNP | MODIFIER | downstream_gene_variant | JA_9 |
|  |  | SNP | MODERATE | missense_variant | JA_2 JA_9 |
|  |  | SNP | MODIFIER | non_coding_transcript_exon_variant | JA_2 JA_9 |
|  |  | SNP | MODERATE | missense_variant | JA_2 JA_9 |
|  |  | SNP | MODERATE | missense_variant | JA_2 JA_9 |
|  |  | SNP | MODERATE | missense_variant | JA_4 JA_11 JA_7 JA_13 |

*: Report of gene in context with JNA.

**Figure S1. Plot showing the Y chromosome mutated genes involved in GO terms of Biological Process.**

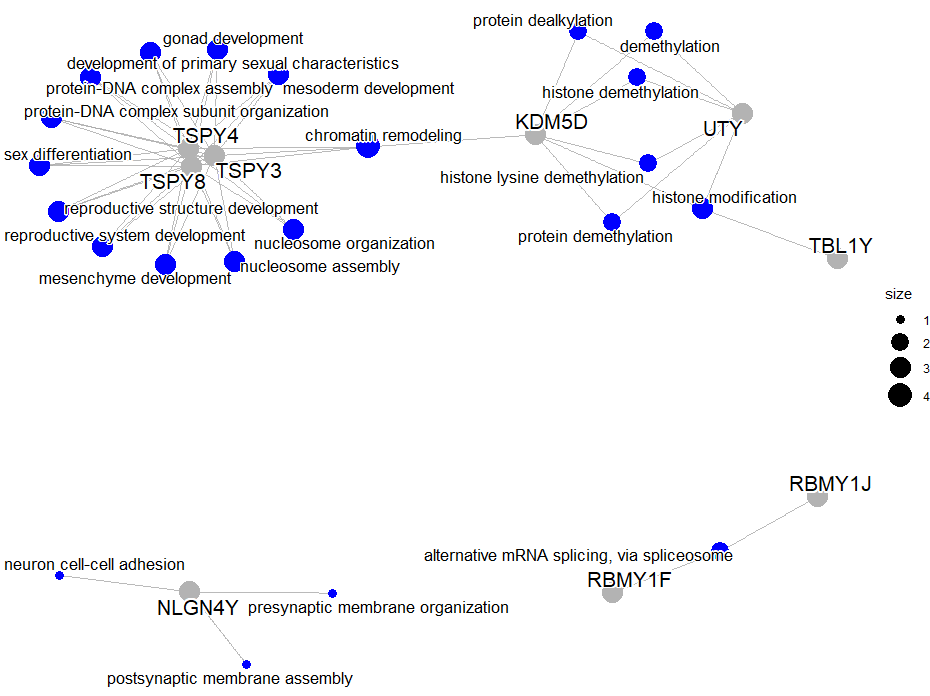

BP and genes are represented by blue and gray circle respectively. Circle size of the BP correspond to number of genes involved.
